# Endocannabinoid levels in patients with Parkinson’s disease with and without levodopa-induced dyskinesias

**DOI:** 10.1101/2020.06.28.20142182

**Authors:** Camila Marchioni, Bruno Lopes Santos-Lobato, Maria Eugênia Costa Queiroz, José Alexandre S. Crippa, Vitor Tumas

## Abstract

**Background:** Levodopa-induced dyskinesias (LID) in Parkinson’s disease (PD) are frequent complications, and the endocannabinoid system has a role on its pathophysiology.

**Objective:** To test the hypothesis that the functioning of the endocannabinoid system would be altered in PD and in LID by measuring plasma and CSF levels of α-N-arachidonoylethanolamine (AEA) and 2-arachidonoyl-glycerol (2-AG) in patients with PD with and without LID and in healthy controls.

**Methods:** Blood and CSF samples were collected from 20 healthy controls, 23 patients with PD without LID, and 24 patients with PD with LID. The levels of AEA and 2-AG were measured using a highly sensitive column switching ultrahigh-performance liquid chromatography-tandem mass spectrometry method.

**Results:** When pooled together, patients with PD had lower plasma and CSF levels of 2-AG and higher CSF levels of AEA compared to healthy controls (Mann-Whitney statistics = 303.0, p = 0.02). Patients with PD without LID had lower CSF levels of 2-AG (Kruskal-Wallis statistics = 7.76, p = 0.02) and higher CSF levels of AEA levels than healthy controls (Kruskal-Wallis statistics = 8.81, p = 0.01).

**Conclusions:** The findings suggest that the endocannabinoid system participates in the pathophysiology of PD symptoms, but its role in the pathophysiology of LID is still unclear.

## Introduction

Parkinson’s disease (PD) is a neurodegenerative condition, and after few years of levodopa therapy motor complications occur, including motor fluctuations and levodopa-induced dyskinesias (LID) [1]. LID have a major impact in the quality of life of patients, especially in the intermediate phase of the disease [2]. LID pathophysiology is not completely understood but is believed to involve the endocannabinoid system (ECS) [3; 4; 5].

The ECS seems to play an important role in the modulation of the basal ganglia [6], mainly through interaction with the dopaminergic transmission based on control of neuronal activity and terminal dopamine release [7; 8]. Anandamide (α-N-arachidonoylethanolamine - AEA) and 2-arachidonoyl-glycerol (2-AG) are the two main neurotransmitters of the ECS, which has two known specific receptors named cannabinoid receptors type 1 (CB1R) and type 2 (CB2R) [6; 9; 10]. There are evidences of the role of the ECS in PD and in LID [7; 11], with increased ECS activity and increased density of CB1R in the basal ganglia of dopamine deficiency animal models of PD, which can be reversed by levodopa therapy [12; 13; 14]. Findings also show that LID are associated with adaptive changes in the functioning of the ECS [7; 15; 16].

Furthermore, few clinical studies reported elevated levels of AEA in the CSF of untreated patients with PD, but no studies have evaluated endocannabinoid levels and their association with LID onset [14; 17]. In this study, we tested the hypothesis that the functioning of the ECS would be altered in PD and in LID by measuring plasma and CSF levels of AEA and 2-AG in patients with PD and healthy controls.

## Materials and methods

### Study design and participants

We conducted an observational cross-sectional study to analyze 2-AG and AEA levels in the plasma and CSF of patients with PD with and without LID and in healthy controls. Patients with PD were recruited from the Movement Disorders Unit of Ribeirão Preto Medical School, Brazil, between October 2015 and May 2017. All patients were diagnosed with PD according to the UK PD Society Brain Bank diagnostic criteria [18]. We invited healthy controls to participate between the companions and caregivers of patients who attended the same hospital. Patients and healthy controls were consecutively invited to participate in the study according to the convenience of using a 1:1 ratio matching for sex and age within four years, and to balance the cases of PD with and without LID. Our goal was to include at least 20 individuals in each group. We defined the presence of LID by a score ≥ 1 in item 4.1 (time spent with dyskinesias) of the International Parkinson and Movement Disorders Society - Unified Parkinson’s Disease Rating Scale (MDS-UPDRS), in addition to clinical confirmation of the presence of LID in the ON-state [19]. Only patients in treatment with levodopa were included.

We excluded participants if they had (1) acute or chronic infections, severe systemic conditions, autoimmune disorders or other neurological diseases; (2) kidney or chronic liver diseases or regular alcohol intake (over 80 g/day for 6 months); and (3) healthy controls if they had neurological or psychiatric disorders. Additionally, we excluded patients with PD if they had (1) dementia according to MDS diagnostic criteria [20]; or (2) mild psychosis defined as a score > 1 on item 1.2 of the MDS-UPDRS Part I (hallucinations and psychosis).

For the analyses, participants were divided into three groups: healthy controls (HC), patients with PD without LID (PD-ND) and patients with PD with LID (PD-D). The study was approved by institutional review board of the Ribeirão Preto Medical School (Number 3.036.243), and each participant provided written informed consent to participate.

### Evaluations

All patients with PD were evaluated by the same movement disorders specialist (B.L.S.L.) using a standardized assessment comprising clinical and epidemiological data, including a definition of levodopa therapy duration and levodopa equivalent daily doses (LEDD), the Hoehn and Yahr stage, the MDS-UPDRS, and the Unified Dyskinesia Rating Scale (UDysRS) [21; 22; 23]. All patients were evaluated in the ON-state. For the UDysRS, we analyzed Part 1B (effect of LID on usual activities described by patients), Part 3 (intensity of LID in seven regions of patient’s body during each of four observed tasks – communication, drinking, dressing and ambulation) and Part 4 (global disability caused by LID during each of these four observed tasks), historical subscore, objective subscore and total UDysRS score. The evaluation sessions of patients with PD with LID were recorded on video for later scoring of the UDysRS objective score.

### Collection, processing and storage of biologic samples

For each participant we collected peripheral blood and CSF on the same day, between 08:00 and 10:00 AM, without fasting. The samples were placed on ice immediately after collection. All patients were instructed to take their regular morning dose of dopaminergic drugs.

Peripheral blood was collected into *Vacutainer* tubes (BD Diagnostics, Plymouth, UK) with anticoagulant (EDTA) by venipuncture, and whole blood was centrifuged at 4°C and 1600g for 15 minutes. Supernatant plasma samples were aliquoted into 1-mL cryotubes, coded, and stored at −80°C until analysis.

CSF was collected through lumbar puncture immediately after the collection of peripheral blood in the lateral recumbent position, using a traumatic Quincke (0.7 mm X 63 mm) needle at the L3/L4 or L4/L5 level. We separated 1-2 mL of CSF for routine analysis (cell counts, glucose, proteins). All remaining CSF (10-12 mL) was collected in polypropylene tubes and gently mixed to avoid gradient effects, centrifuged at 4°C at 4000g for 10 minutes to remove cells, aliquoted into 1-mL cryotubes, coded, and stored at −80°C until use without preservatives added. CSF samples contaminated with blood (CSF red cells > 500 per mm3) were excluded.

### Lipid extraction and UHPLC-MS/MS conditions

For lipid extraction, CSF was mixed with acetone (2:1, v/v) in the presence of AEA-d4 and 2-AG-d5 as internal standards. The supernatant was mixed with toluene for liquid-liquid extraction and the organic phase was dried and analyzed [24; 25]. This procedure was based on the method described previously [26]. For plasma samples, the procedure was a simple precipitation of proteins with acetonitrile, as described elsewhere [24].

UHPLC-MS/MS analysis was performed with a two-dimensional Waters® UHPLC-MS/MS system (Waters Corporation, Milford, MA, USA) coupled with a Xevo® TQ-D triple-quadrupole operating in positive electrospray ionization (ESI+) mode with multiple reaction monitoring (MRM). Plasma samples (100 µL) were injected into a first RP-8 ADS column (25 mm × 4 mm × 25 μm) and then into a Kinetex C18 core-shell column (100 mm × 2.1 mm × 1.7 μm) in the second dimension [24]. RP-8 ADS was used for enrichment with traces of endocannabinoids and macromolecular matrix size exclusion, while the core-shell column was used for chromatographic separation. The prepared CSF samples (10 µL) were injected directed into the second column (Kinetex C18 core-shell) [24].

### Statistical analysis

To compare clinical and epidemiological data between groups, we performed the Mann-Whitney and Kruskal-Wallis tests followed by Dunn’s test for multiple comparisons for continuous variables, and the chi-square test for categorical variables. To compare the levels of endocannabinoids between groups, we used the Mann-Whitney and Kruskal-Wallis tests followed by Dunn’s test for multiple comparisons. To compare two continuous variables, we performed Spearman’s Rho correlation test. All analyses were performed using SPSS for Windows version 23.0 (SPSS Inc., Chicago, USA) and figures were created using *GraphPrism* for Windows version 5.0 (GraphPad Software Inc., La Jolla, USA).

## Results

### Clinical and demographic data

Sixty-seven participants fulfilled the inclusion and exclusion criteria. We were unable to collect CSF from four patients with PD (one with LID and three without LID) due to technical problems during the lumbar puncture. The clinical and demographic data of participants are summarized in Table 1.

**Table 1.** Clinical and epidemiological data of patients with Parkinson’s disease and healthy controls.

| General characteristics | HC (n=20) | PD-ND (n=20) | PD-D (n=23) | p-value |
| --- | --- | --- | --- | --- |
| Male sex, % (n) | 30 (6) | 75 (15) | 65.2 (15) | 0.002<sup>a</sup> |
| Age at the time of evaluation<sup>b</sup> | 63 (61-67) | 62 (53-66) | 59 (53-64) | 0.06<sup>c</sup> |
| Weight (kg) | NA | 72.5 (66-79) | 73 (60-83) | 0.63<sup>d</sup> |
| Age at onset of PD<sup>b</sup> | NA | 54.5 (48-61) | 47 (42-51) | 0.03<sup>d</sup> |
| Disease duration (years)<sup>b</sup> | NA | 8 (5-12) | 11 (8-16) | 0.01<sup>d</sup> |
| Levodopa therapy duration (years)<sup>b</sup> | NA | 5 (3-8) | 8 (6-12) | 0.05<sup>d</sup> |
| Levodopa dose at evaluation (mg/day)<sup>b</sup> | NA | 500 (400-850) | 850 (750-1200) | 0.003<sup>d</sup> |
| Levodopa equivalent daily dose (mg/day)<sup>b</sup> | NA | 950 (500-1275) | 1400 (1090-2050) | 0.002<sup>d</sup> |
| Tremor as initial motor symptom, % (n) | NA | 70 (14) | 43.5 (10) | 0.12<sup>a</sup> |
| Tremor dominant phenotype at evaluation, % (n) | NA | 80 (16) | 26.1 (6) | 0.002<sup>a</sup> |
| MDS-UPDRS Part I<sup>b</sup> | NA | 10.5 (3-17) | 10 (8-17) | 0.28<sup>d</sup> |
| MDS-UPDRS Part II<sup>b</sup> | NA | 15 (5-18) | 19 (12-25) | 0.02<sup>d</sup> |
| MDS-UPDRS Part III<sup>b</sup> | NA | 30.5 (25-40) | 25 (19-34) | 0.3<sup>d</sup> |
| MDS-UPDRS Part IV<sup>b</sup> | NA | 0 (0-5) | 11 (8-15) | < 0.001<sup>d</sup> |
| MDS-UPDRS Total score<sup>b</sup> | NA | 57.5 (45-73) | 64 (57-88) | 0.04<sup>d</sup> |
| Hoehn & Yahr stage<sup>b</sup> | NA | 2 (2-2) | 2 (2-3) | 0.37<sup>d</sup> |
| UDysRS Part 1B<sup>b</sup> | NA | NA | 21 (16-25) | - |
| UDysRS Part 3<sup>b</sup> | NA | NA | 14 (11-19) | - |
| UDysRS Part 4<sup>b</sup> | NA | NA | 7 (4-9) | - |
| UDysRS Historical subscore<sup>b</sup> | NA | NA | 28 (17-39) | - |
| UDysRS Objective subscore<sup>b</sup> | NA | NA | 20 (15-27) | - |
| UDysRS Total score<sup>b</sup> | NA | NA | 53 (36-62) | - |

<sup>a</sup>Chi-square test comparing frequencies between healthy controls, patients with PD with and without levodopa-induced dyskinesias
<sup>b</sup>Values in median (interquartile range)
<sup>c</sup>Kruskal-Wallis test comparing medians between healthy controls, patients with PD with and without levodopa-induced dyskinesias
<sup>d</sup>Mann-Whitney test comparing medians between patients with and without levodopa-induced dyskinesias
Abbreviations: HC, healthy controls; NA, not applied; PD-D, Patients with PD with levodopa-induced dyskinesias; PD-ND, Patients with PD without levodopa-induced dyskinesias.

Men predominated in the PD-ND (62.5%) and PD-D (78.3%) groups, and women in the HC group (70%). There were no significant age differences across groups. Patients with PD with LID had an earlier onset of disease, longer disease duration, longer levodopa therapy, and higher LEDD intake compared to patients with PD with LID patients.

### Plasma and CSF AEA and 2-AG levels

The endocannabinoid levels of 2-AG and AEA in the plasma and CSF samples of patients with PD and healthy controls are presented in Figure 1 and Table 2.

**Table 2.** Endocannabinoid plasma and CSF levels in patients with Parkinson’s disease and healthy controls.

| Endocannabinoid | HC | PD-ND | PD-D | p-value |
| --- | --- | --- | --- | --- |
| Plasma |  |  |  |  |
| n | 20 | 23 | 24 |  |
| 2-AG, ng/mL | 0.57<br>(0.24-0.67) | 0.26<br>(0.11-0.55) | 0.21<br>(0.08-0.78) | 0.07 |
| AEA, ng/mL | 0.19<br>(0.16-0.3) | 0.26<br>(0.16-0.47) | 0.19<br>(0.14-0.24) | 0.33 |
| CSF |  |  |  |  |
| n | 20 | 20 | 23 |  |
| 2-AG, ng/mL | 0.41<br>(0.27-0.63) | 0.25*<br>(0.19-0.34) | 0.29<br>(0.23-0.64) | 0.02 |
| AEA, ng/mL | 0.64<br>(0.59-0.82) | 0.85*<br>(0.75-0.95) | 0.81<br>(0.62-0.97) | 0.01 |

Values are expressed in medians (interquartile range). Kruskal-Wallis test was used to compare medians between healthy controls, patients with PD with and without levodopa-induced dyskinesias.
\*Statistically significant differences relative to the HC group
Abbreviations: 2-AG = 2-arachidonoyl-glycerol; AEA = anandamide; HC = healthy controls; PD-D = Patients with PD with levodopa-induced dyskinesias; PD-ND = Patients with PD without levodopa-induced dyskinesias.

**Figure 1.**
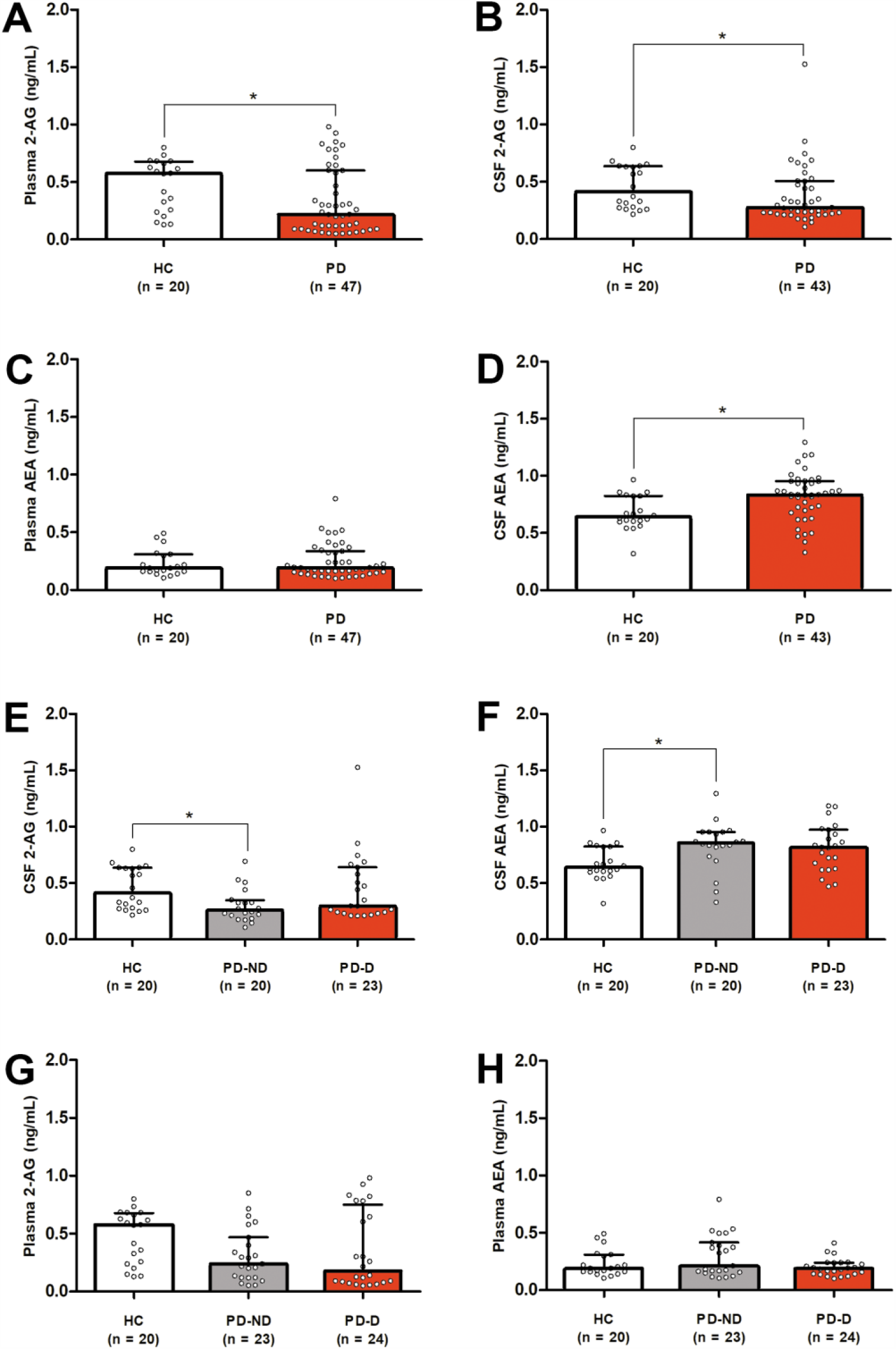
Endocannabinoid plasma and CSF levels in patients with Parkinson’s disease and healthy controls. A: Plasma 2-AG in HC, PD-ND and PD; B: Plasma AEA in HC, PD-ND and PD; C: CSF 2-AG in HC, PD-ND and PD; D: CSF AEA in HC, PD-ND and PD; E: Plasma 2-AG in HC and all patients with PD; F: Plasma AEA in HC and all patients with PD; G: CSF 2-AG in HC and all patients with PD; H: CSF AEA in HC and all patients with PD. Abbreviations: 2-AG = 2-arachidonoyl-glycerol; AEA = anandamide; HC = healthy controls; PD-D = Patients with PD with levodopa-induced dyskinesias; PD-ND = Patients with PD without levodopa-induced dyskinesias.

The comparative analysis of the groups of patients with PD and HC showed that patients with PD had lower plasma and CSF levels of 2-AG (plasma: Mann-Whitney statistics = 303.0, p = 0.02; CSF: Mann-Whitney statistics = 286.0, p = 0.03) (Figure 1A and 1B), and higher CSF levels of AEA than HC (plasma: Mann-Whitney statistics = 460.5, p = 0.9; CSF: Mann-Whitney statistics = 234.0, p = 0.003) (Figures 1C and 1D). The comparative analysis considering the presence of LID showed that patients of the PD-ND group had lower CSF levels of 2-AG and higher CSF levels of AEA than HC (2-AG: Kruskal-Wallis statistics = 7.76, p = 0.02, difference in rank sum = −15.95, p < 0.05; AE: Kruskal-Wallis statistics = 8.81, p = 0.01, difference in rank sum = 16.35, p < 0.05) (Figures 1E and 1F). There were no differences between groups in the plasma levels of endocannabinoids (2-AG: Kruskal-Wallis statistics = 5.24, p = 0.07; AEA: Kruskal-Wallis statistics = 2.16, p = 0.33) (Figure 1G and 1H).

### Correlations between endocannabinoid levels and clinical variables

In respect to endocannabinoid levels in the CSF, body weight was associated with AEA levels (Spearman’s ρ = −0.41, p = 0.006, n = 43) and MDS-UPDRS Part 1 score was associated with AEA and 2-AG and AEA levels (2-AG: Spearman’s ρ = 0.35, p = 0.02, n = 43; AEA: Spearman’s ρ = −0.34, p = 0.02, n = 43). Regarding plasma, advanced Hoehn & Yahr stages were associated with lower AEA levels (Spearman’s ρ = −0.36, p = 0.01, n = 47), but increased time spent with dyskinesias (item 4.1 of the MDS-UPDRS) in PD patients with LID was associated with higher AEA levels (Spearman’s ρ = 0.45, p = 0.02, n = 24).

There were no correlations between the levels of endocannabinoids in plasma or CSF and age at onset of PD, disease duration, levodopa therapy duration and dose at evaluation, LEDD, and MDS-UPDRS total and sub-item scores, initial motor symptoms, and clinical motor phenotype. Also, there were no correlations between endocannabinoid levels and UDysRS scores.

When patients and controls were pooled together, we found no correlations between plasma levels of AEA and 2-AG (Spearman’s ρ = 0.156, p = 0.206, n = 67), but CSF levels of AEA and 2-AG tend to be in opposite directions (Spearman’s ρ = −0.221, p = 0.08, n = 63). Plasma and CSF levels of 2-AG had inverse patterns in their levels (Spearman’s ρ = −0.295, p = 0.01, n = 63).

Longer intervals between sample collection and freezing were associated with higher plasma 2-AG levels (Spearman’s ρ = 0.46, p = < 0.001, n = 61) (Figure 2A), longer plasma storage times were associated with lower plasma AEA levels (Spearman’s ρ = - 0.52, p = < 0.001, n = 67), and longer CSF storage times were associated with higher CSF 2-AG levels (Spearman’s ρ = 0.33, p = 0.007, n = 63) (Figure 2B). Covariance analyses did not show a significant effect of these variables on the measurements of endocannabinoid levels in the three groups. There was no correlation between CSF 2- AG and AEA levels and cell counts, proteins, glucose, and chloride levels in CSF.

**Figure 2.**
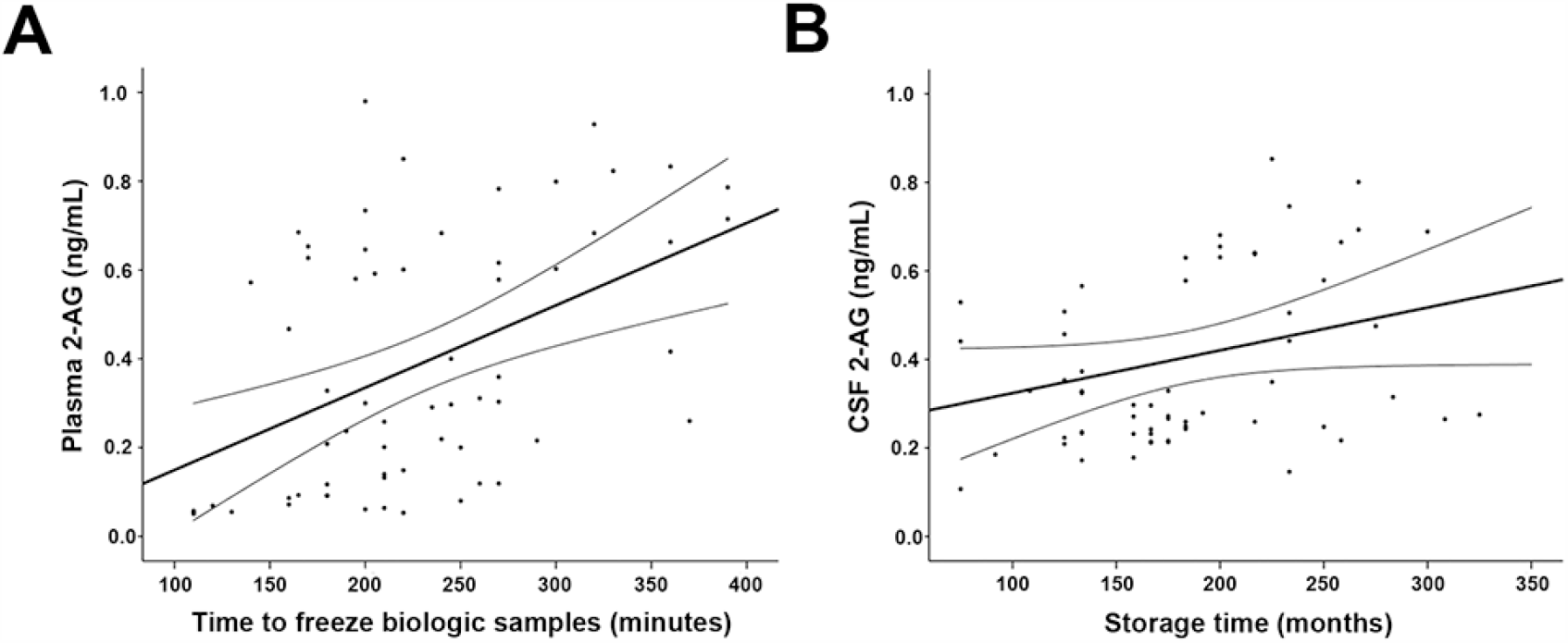
Correlations between plasma and CSF 2-AG levels and sample processing intervals. A: Correlation between plasma 2-AG levels and time to freeze biologic samples in minutes; B: Correlation between CSF 2-AG levels and storage time in months. Abbreviations: 2-AG = 2-arachidonoyl-glycerol.

## Discussion

In our study, we found elevated CSF levels of AEA and decreased CSF and plasma levels of 2-AG in patients with PD, in comparison to healthy controls. These CSF alterations in endocannabinoid levels were observed only in patients with PD without LID, but not in those with LID.

There are few clinical studies on CSF AEA levels in patients with PD. The first study reported higher levels of AEA in the CSF of patients with PD compared to healthy controls [17]. Later, the same group demonstrated that CSF levels of AEA were elevated in patients with PD without dopaminergic therapy, and that chronic dopaminergic replacement restored CSF levels of AEA to normal levels [16]. The authors suggested that elevated CSF AEA levels would be part of the compensatory mechanisms induced by striatal dopaminergic loss, and that treatment with dopaminergic drugs would restore AEA levels to normal [16; 17; 27]. No previous studies analyzed 2-AG CSF levels in patients with PD. Our findings are consonant with these original observations showing increased CSF AEA levels in patients with PD, but additionally revealed that 2-AG levels are reduced in the CSF of these patients. This shows that the CSF levels of the two major endocannabinoids follow an inverse relationship in patients with PD.

Many experimental observations suggested that the functioning of the ECS is altered in PD. Preliminary studies reported alterations in endocannabinoid levels in the basal ganglia in animal models of PD [7; 13]. There were increased levels of endocannabinoids in basal ganglia of rats treated with reserpine, but especially noted a seven-fold elevation in the levels of 2-AG in the globus pallidus. They suggested that, in the parkinsonian state, increased levels of 2-AG in the globus pallidus would be a consequence of increased activity in the indirect pathway [13]. Also, elevated levels of AEA and 2-AG in the basal ganglia of MPTP-lesioned primates were described [7]. In both studies, treatment with dopaminergic drugs promoted a significant reduction in the levels of endocannabinoids in these basal ganglia, and in one of them it was accompanied by increased locomotion of the reserpine-treated rats [7; 13]. These observations suggest that changes in the ECS might result from the development of compensatory modifications induced by a reduction in striatal dopamine levels, which would be characterized by increases in the levels of endocannabinoids in the basal ganglia. The findings available also indicate that dopamine play a modulatory role in the ECS activity and that dopaminergic therapy in PD would restore the normal functioning of this system and reduce the abnormally elevated endocannabinoid levels [7; 13].

However, other experimental studies suggested that changes in ECS function in the basal ganglia following dopaminergic striatal denervation seem to be more complex. One study using an MPTP mouse model of PD showed that, following MPTP administration, the levels of 2-AG in the ventral midbrain increased signi?cantly for two days after administration; however, these levels were almost back to control levels seven days after treatment. On the other hand, there was a slow decline in the level of 2-AG in the striatum, reaching significantly lower values at seven days until 21 days after treatment with MPTP [28]. In another study using unilaterally 6-OHDA-lesioned rats, AEA levels reduced considerably in the caudate-putamen ipsilateral to the lesion; however, neither acute nor chronic levodopa treatment affected the levels of endocannabinoids in these animals [16].

Our findings agree with studies that reported increased levels of AEA in the CSF/brain of patients and animal models of PD, and with experimental studies that reported decreased levels of 2-AG [7; 13; 17; 28]. We found increased CSF AEA levels in PD patients using dopaminergic therapy. As levodopa treatment has been reported to reduce CSF AEA levels back to normal, we expected that endocannabinoid levels were influenced by the intake of dopaminergic drugs in our study. Unfortunately, we could not explore this association, as a drug-näive group of patients with PD was not included, as our aim was to verify changes in the ECS associated with the development of LID. Therefore, we cannot rule out the possibility that endocannabinoid levels could be different if patients were not on dopaminergic drug therapy.

The reasons for the differences observed in the endocannabinoid levels of our patients with and without LID are unclear. PD-D group presented higher LEDD intake and longer levodopa therapy than PD-ND group; however, we found no significant correlations between endocannabinoid levels and LEDD intake and duration of levodopa therapy. Based on these findings, we are unable to conclude whether endocannabinoid levels vary according to the dose and duration of dopaminergic therapy.

Another interesting finding was that changes in AEA and 2-AG levels in the CSF of patients with PD tend to take opposite directions. There is evidence that AEA inhibits the metabolism and physiological actions of 2-AG in the striatum, which could explain these findings [29; 30]. However, we found no significant correlations between AEA and 2-AG levels in the CSF as could be expected based on this hypothesis.

In respect to plasma, we found no significant alterations in the levels of endocannabinoids across groups, and there were no essential correlations between plasma and CSF endocannabinoid levels; therefore, it is possible to assume that the change in plasma endocannabinoid levels was presumably of central origin.

Our findings support the hypothesis that the physiology of the ECS is altered in PD. However, the association between the human plasma and CSF endocannabinoid levels and LID in PD is still unclear. The role of the ECS in the pathophysiology of LID seems to be complex, since endocannabinoids act in retrograde signaling in many circuits of the basal ganglia and could modulate different neurotransmitter systems as glutamate, adenosine, but especially dopamine transmission [31].

Our study has limitations, among which we highlight the low sample size and the absence of a drug-näive group of patients with PD, which would allow us to investigate variations in the endocannabinoid levels of untreated patients. Together, these findings suggest that the endocannabinoid system participates in the pathophysiology of PD symptoms, but its role in the pathophysiology of LID is still unclear.

## Data Availability

The datasets generated during and/or analyzed during the current study are available from the corresponding author on reasonable request.

## Acknowledgments

We would like to thank Ângela Vieira Pimentel, Larissa Serveli, Manuelina Macruz Capelari and Nathália Novaretti (Ribeirão Preto Medical School, University of São Paulo) for technical support.

## Conflict of Interest

Dr. CRIPPA is co-inventor (Mechoulam R, Zuardi AW, Kapczinski F, Hallak JEC, Guimarães FS, Crippa JA, Breuer A) of the patent ‘Fluorinated CBD compounds, compositions and uses thereof. Pub. No.: WO/2014/108899. International Application No.: PCT/IL2014/050023’, Def. US no. Reg. 62193296; 29/07/2015; INPI on 19/08/2015 (BR1120150164927). The University of São Paulo has licensed the patent to Phytecs Pharm (USP Resolution No. 15.1.130002.1.1). The University of São Paulo has an agreement with Prati-Donaduzzi (Toledo, Brazil) to ‘develop a pharmaceutical product containing synthetic cannabidiol and prove its safety and therapeutic efficacy in the treatment of epilepsy, schizophrenia, Parkinson’s disease, and anxiety disorders’. Dr. CRIPPA has received travel support from BSPG-Pharm and is a medical advisor of SCBD Centre. Dr. CRIPPA has a grant from University Global Partnership Network (UGPN)–Global priorities in cannabinoid research excellence. Dr. CRIPPA is a member of the international advisory board of the Australian Centre for Cannabinoid Clinical and Research Excellence (ACRE), funded by the National Health and Medical Research Council through the Centre of Research Excellence. Dr. CRIPPA is recipient of CNPq 1A productivity fellowship.

## Notes

### Competing Interest Statement

The authors have declared no competing interest.

### Funding Statement

This study was supported by the Fundação de Amparo à Pesquisa do Estado de São Paulo (FAPESP; 159688/2015-9) and Conselho Nacional de Desenvolvimento Científico e Tecnológico (CNPq; 2012/17626-7).

### Author Declarations

The study was approved by institutional review board of the Ribeirão Preto Medical School (Number 3.036.243), and each participant provided written informed consent to participate.

